# Indirect costs and scientific impact at NIMH

**DOI:** 10.1101/2024.09.23.24314209

**Authors:** Roy H. Perlis

## Abstract

The National Institutes of Health (NIH) awards additional funds for extramural research to support research infrastructure and administration, such that the total cost of a given research project depends on where it is conducted. We sought to understand whether greater indirects were associated with a greater scientific impact of NIH-funded work. The NIH RePORTER database was queried to retrieve all R01, R21, or R03-funded research proposals for which NIMH was listed as the primary funding source for proposals funded between 2012 and 2023. We applied multiple regression to examine the association between indirect rate and measures of scientific impact, including number of publications, their citation impact in terms of H-index per grant and total citations, and the number of patents associated with each grant. Of 5,143 projects, reflecting $9.85 billion, mean indirect rate was 47.9% (SD 16.2%). Greater indirect rate was associated with modest but statistically significantly greater number of publications (+0.30 per 10% increase in indirect rate, 95% CI 0.08-0.51); H-index at 5 years (+0.25 per 10% increase in indirect rate, 95% CI 0.18-0.33; Figure 1); and total citations (+29.71 per 10% increase in indirect rate, 95% CI 17.86-41.57). Each 10% increase in indirect rate was associated with a 20% increase in odds of patent filing (aOR 1.20, 95% CI 1.05-1.37). The results suggest small incremental benefits from conduct of research at higher-cost institutions and provide data for policymakers to consider in weighing the costs against potential benefits of work at such institutions.

## Introduction

For every dollar paid for extramural research, the National Institutes of Health (NIH) provide additional funds to support research infrastructure and administration (F&A) – so-called indirect charges. The indirect rate for a given institution is determined by the NIH Division of Financial Advisory Services, on the basis of materials submitted by that institution to document its costs. A decade ago, a *Nature* investigation^1^ found that these rates varied widely, from 20-85%, following efforts to ensure that indirect funds were no longer used for expenses like yacht depreciation or home furnishings for institutional leadership. At that time, they accounted for around a fifth of NIH’s total budget^2^.

In practice, this variation means that the same research project conducted at two institutions could have markedly different total costs to the federal government – and, ultimately, to the public. This study investigated whether this additional investment yielded additional benefit, using data from the National Institute of Mental Health (NIMH). That is, to what extent did more expensive research at NIMH associate with greater quantifiable scientific impact, measured by publications, citations, and patent filings.

## Methods

The author queried the NIH RePORTER database to retrieve all research proposals categorized as R01, R21, or R03 between 2012-2023 with NIMH as the primary funding source. Total direct-cost investment in each project was determined by summing over all project years; indirect rate was calculated as the ratio between total indirect and total direct cost. Dollar values were inflation-adjusted to 2023 values^3^.

The first outcome examined was output of each project in publications, using Pubmed ID’s (PMID) listed in RePORTER. The second was citation impact, in terms of H-index^4^ per grant, reflecting the number of papers associated with a given grant each cited at least that number of times. The European Pubmed Central API was queried to determine the number of publications citing a given PMID each year, limited to 5 years after initial publication; sensitivity analysis examined estimates for grants funded prior to 2019, and total number of citations. The third outcome was number of patents for each grant based on RePORTER data; values were dichotomized to indicate presence or absence of at least one patent based on visual inspection of distribution.

### Analysis

Multiple regression (linear for publications and citations, logistic for patents) was applied to examine the association between calculated indirect rate and measures of scientific impact, adjusted for grant type (R01, R21, or R03), funding year, number of investigators, and total funding. Analyses used R 4.3.2 with two-tailed p-values <.05 representing statistical significance. This research was not evaluated by an institutional review board.

## Results

Of 5,143 R01, R03, and R21 projects initially funded between 2012 and 2023, reflecting $9.85 billion, mean indirect rate was 47.9% (SD 16.2%). In linear regression, publications were significantly but modestly greater (+0.30 per 10% increase in indirect rate, 95% CI 0.08-0.51); H-index at 5 years was likewise significantly but modestly greater (+0.25 per 10% increase in indirect rate, 95% CI 0.18-0.33; Figure 1), as were total citations (+29.71 per 10% increase in indirect rate, 95% CI 17.86-41.57;). In logistic regression, each 10% increase in indirect rate was associated with a 20% increase in odds of patent filing (aOR 1.20, 95% CI 1.05-1.37).

**Figure 1.**
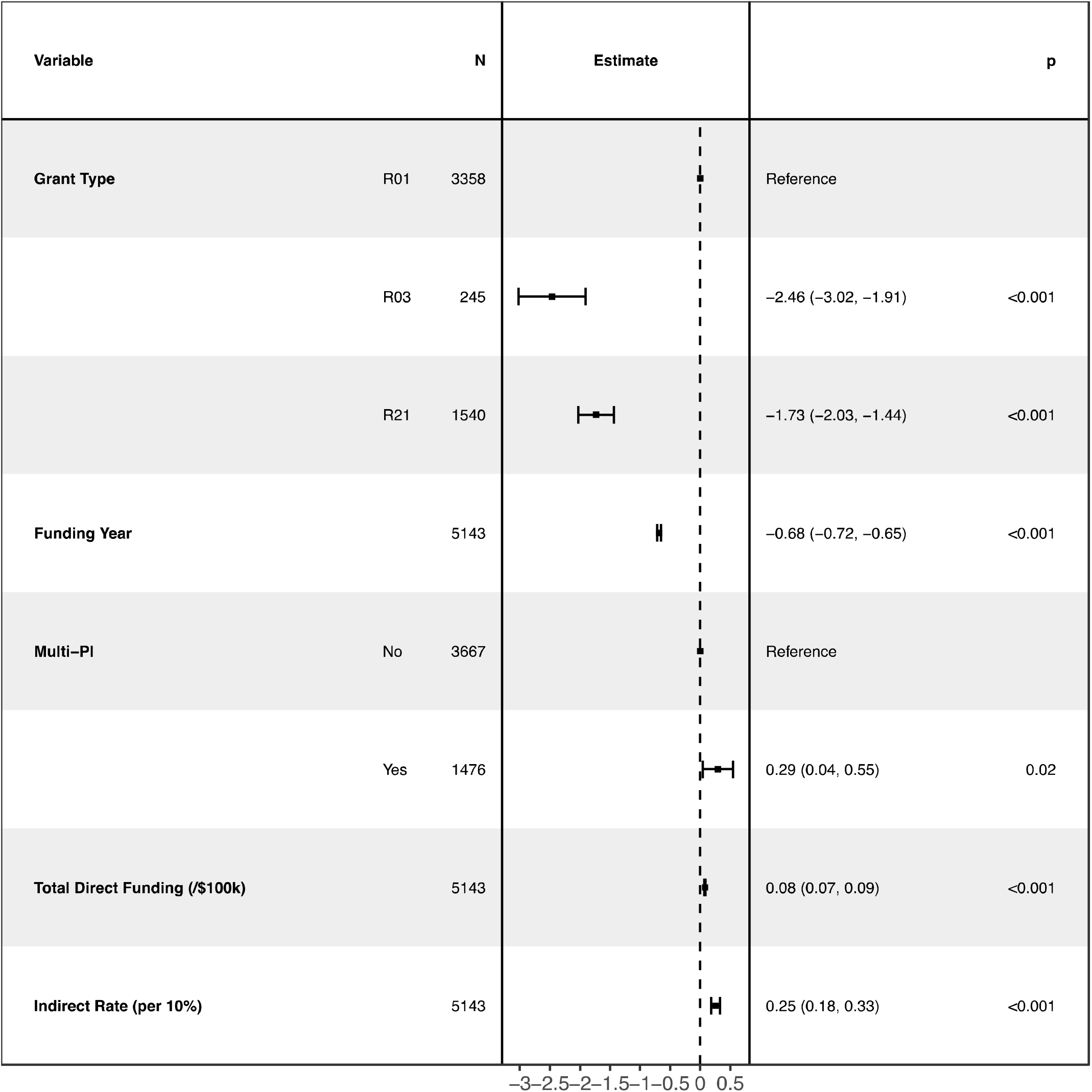
Multiple linear regression model of association between indirect rate and H-index at five years

## Discussion

In this analysis of $9.85 billion in NIMH R01, R03, and R21 grants funded between 2012-2023, greater indirect rates were associated with modest increases in multiple metrics of scientific productivity. Unlike other metrics, the value of a given unit change may be difficult to interpret, particularly for publications and citation. However, a reasonable question for policymakers is whether science should cost different amounts depending on where it is conducted – and, if so, how much each incremental patent or publication should cost. Indeed, in many other countries, indirect rates are constant across institutions^1^.

We note multiple limitations. First, these associations reflect a single NIH institute. We could identify only one prior study using NIMH grant-level data, to examine expenditure on child psychiatry^5^. This institute was selected simply because it represents the author’s primary funding source, and estimating all outputs was too computationally costly to examine every institute. However, future work could examine other institutes and federal grant funding more generally. Second, this report focuses on a narrow set of scientific productivity measures, while biomedical research may contribute in numerous other ways. Still, the measures employed represent standard and widely-used academic metrics, while other aspects of productivity likely introduce greater subjectivity.

Perhaps most importantly, in identifying a cross-sectional association, we cannot assume that this relationship is causal, nor determine a mechanism by which greater indirect costs could increase productivity. Higher-cost institutions may attract researchers with greater productivity; conversely, such institutions may be better able to promote the work of their researchers.

Despite these limitations, our results suggest the incremental benefit of research at more costly institutions is likely to be modest. In an era when much work can be done remotely, efforts to shift some scientific work to institutions with lower indirect costs merit consideration. In other words, policymakers should consider whether a cost-aware strategy might allow more productive deployment of the fixed (and, in inflation-adjusted terms, shrinking) budget available to them.

## Data Availability

All data used is publicly available via the NIH RePORTER Database.

https://reporter.nih.gov/

## Data Availability

All data used is publicly available via the <u>NIH RePORTER Database</u>.

## Competing Interests

Dr. Perlis has received fees for scientific advising from Belle AI, Genomind, Circular Genomics, Mila Health, Vault Health, and Alkermes, outside of the present work. He holds equity in Psy Therapeutics and Circular Genomics. He serves as Editor-in-Chief of *JAMA Artificial Intelligence* and Associate Editor at *JAMA Network – Open*.

## Funding

Dr. Perlis is supported in part by the National Institute of Mental Health (R01MH123804, U01MH136059). The sponsors did not contribute to the design and conduct of the study; collection, management, analysis, and interpretation of the data; preparation, review, or approval of the manuscript; and decision to submit the manuscript for publication. Dr. Perlis had full access to all the data in the study and takes responsibility for the integrity of the data and the accuracy of the data analysis.

